# An integrative machine learning approach to discovering multi-level molecular mechanisms of obesity using data from monozygotic twin pairs

**DOI:** 10.1101/2019.12.19.19015347

**Authors:** Milla Kibble, Suleiman A. Khan, Muhammad Ammad-ud-din, Sailalitha Bollepalli, Teemu Palviainen, Jaakko Kaprio, Kirsi H. Pietiläinen, Miina Ollikainen

## Abstract

We combined clinical, cytokine, genomic, methylation and dietary data from 43 young adult monozygotic twin pairs (aged 22 – 36, 53% female), where 25 of the twin pairs were substantially weight discordant (delta BMI > 3kg/ m^2^). These measurements were originally taken as part of the TwinFat study, a substudy of The Finnish Twin Cohort study. These five large multivariate data sets (comprising 42, 71, 1587, 1605 and 63 variables, respectively) were jointly analysed using an integrative machine learning method called Group Factor Analysis (GFA) to offer new hypotheses into the multi-molecular-level interactions associated with the development of obesity. New potential links between cytokines and weight gain are identified, as well as associations between dietary, inflammatory and epigenetic factors. This encouraging case study aims to enthuse the research community to boldly attempt new machine learning approaches which have the potential to yield novel and unintuitive hypotheses. The source code of the GFA method is publically available as the R package GFA.

## 1 Introduction

Worldwide, obesity has nearly tripled since 1975, according to the World Health Organisation. In 2016, more than 1.9 billion adults were over-weight, of whom over 650 million were obese (1). A raised Body Mass Index (BMI), which is a common measure used to define obesity, is a major risk factor for non-communicable diseases such as cardiovascular diseases, type 2 diabetes mellitus (T2DM), chronic kidney disease, musculoskeletal disorders (especially osteoarthritis) and certain cancers (2-4). There is evidence that high BMI is really driving the unfavourable changes in disease associated biomarkers (5). Recently, Romieu et al (6) reviewed the evidence of the associations between energy balance and obesity and concluded that the main driver of weight gain is energy intake that exceeds its expenditure. So excessive energy intake which is not compensated by energy expenditure leads to excess weight gain, which over time can lead to a multitude of chronic health problems.

However, the weight gain trajectory is far from being this straightforward. Key players likely include genetics, epigenetics, type and quality of diet, exercise and lifestyle, microbiome composition and hormonal effects as well as medications and societal and psychological factors, all interacting in such a complex manner that often researchers can focus on only one or two of these features at a time (7). For example, there are studies highlighting the heritability of obesity (8; 9), estimates of which range between 40 and 70%. Indeed monozygotic (MZ) twins are very rarely BMI discordant, i.e. differ significantly in weight. The first genetic locus to show robust association with BMI and obesity risk, namely FTO, was discovered via a genome-wide association study (GWAS) in 2007 (10; 11) and since then over 500 genetic loci have been found, amongst other things providing strong support for a role of the central nervous system in obesity susceptibility (8). It is now clear that most cases of obesity are polygenic and multifactorial and the full picture of how genetic and other factors jointly influence at a molecular level individual preferences for and responses to diet and physical activity remains largely beyond our comprehension.

With the advent of large and diverse data sets coupled with innovative computational tools which can look at these data sets in combination, this Holy Grail may be getting a step closer. Here we present one example of Big Data being combined with machine learning in an innovative way to infer a multilayer picture of the mechanisms behind obesity. What we present is not a finished answer, but an encouraging case study which aims to enthuse the research community to boldly attempt new machine learning approaches which have the potential to yield novel and unintuitive hypotheses.

In this paper, we combine genetic, methylation, clinical, cytokine, dietary and lifestyle data from MZ twin pairs, many of whom are discordant for obesity – a rare data set - which is then input into a machine learning tool never before used on twin data to elucidate interactions at multiple molecular levels in the development of obesity. The outcome results encouragingly highlight many known associations as well as suggesting novel links that could offer new hypotheses of molecular mechanisms. We start with a brief background to twin studies to highlight the novelty of our method in this domain. We then describe the method in accessible terms for an interdisciplinary audience and only then proceed to the key results on obesity. Finally, we offer our thoughts on future possibilities of such an approach.

### 1.1 Twin studies

So-called *classical twin methods* have focused on estimating the heritability of different phenotypes by comparing occurrences of the phenotype in those monozygotic (MZ) and dizygotic (DZ) twin pairs where at least one twin exhibits the phenotype of interest. The basic premise is that if genetics influences a particular phenotype, then the occurrence of that phenotype for both twins will be more common within MZ twin pairs, who share their whole genomic sequence, than DZ twin pairs, who share roughly 50% of their segregating genes akin to non-twin siblings (12). Such heritability studies using large twin repositories cover the whole range of complex phenotypes (13; 14) including obesity (8) and many obesity-related phenotypes. For example, by looking at 1,126 twin pairs, Goodrich et al (15) identified heritable bacterial taxa of the gut microbiome. The most heritable taxa was the family Christensenellaceae (phylum Firmicutes) which is enriched in lean individuals and has been shown to limit adiposity gain when given as faecal transplants to mice deficient in the taxa, suggesting that heritable microbes could influence adiposity.

Another popular twin design is the so-called *co-twin control method* whereby within-pair comparisons of trait discordant MZ twin pairs are used to identify factors associated with the trait, against a background of equivalent age, sex and genotype (which are perfectly matched for the twins) and in part also ruling out the influence of environment (which is partially matched, and termed as shared environment) (16). The first such studies helped prove the effect of smoking on lung cancer (17). Illustrative examples relevant to obesity would be the recent study in BMI-discordant MZ twin pairs revealing sub-types of obesity based on both clinical traits and gene expression in subcutaneous adipose tissue (18), as well as that based on DNA methylation in leucocytes (19). Such studies usually employ classical machine learning approaches such as linear mixed-effects models.

For a thorough exposition of twin studies, the reader is referred to the review article of van Dongen et al. (13). In the current article, we also look at MZ twin pairs discordant for obesity and in particular use machine learning methods to search for associations in any differences between the heavier and leaner individuals in such pairs. Such associations including dietary and lifestyle data could give hints as to behavioural mechanisms for the twins differing in BMI despite sharing the same genotype, whereas associations involving the other types of data could highlight consequences and/or causes of obesity which are genotype independent. To our knowledge, machine learning techniques of the form proposed here have not previously been applied as part of twin studies.

### 1.2 Machine learning

Machine learning and artificial intelligence (AI) have been applied in the field of medicine for over a decade and, with the now routine collection of large data sets in all research fields and the affordability of computing power, the number of application areas is rising sharply. Although to a lesser extent than in the drug discovery sector, machine learning is also being used in innovative ways in the area of disease prevention (see the discussion in (20)) and it is here that our current contribution sits.

Machine learning refers to mathematical algorithms that have been coded into computer programs. Their function is to look at the data set of interest and “learn” patterns in that data and possibly also predict data values not available to the researcher. Most algorithms achieve this by having an underlying mathematical model to describe the type of data defined by a set of unknown parameters and then calculating the parameters that best fit the particular data set. Camacho et al. have recently written a very clear introduction to machine learning in biological applications (21), including definitions of the most commonly used terms and examples of recent developments.

Here we use an advanced machine learning method called Group Factor Analysis (GFA); see the **Methods** section. It takes as its input multiple data sets (or matrices), where each matrix has samples in its rows and variables in its columns. The idea is to have variables of a similar type in each matrix and in the current work we have five matrices, one each for clinical, cytokine, genotype, methylation and dietary data.

Our samples correspond to twin pairs and the values in the matrices correspond to the difference in measurement of the variable for the twins in the pair (we always subtract the value for the leaner twin from the value for the heavier twin). GFA then learns associations between variables within data sets and between data sets, finding associations of the form “heavier twins who consume a lot more of nutrient x than their leaner twin, tend to have higher levels of cytokine y than their leaner twin and greater methylation at CpG sites z and w”.

The output of GFA can be viewed via so-called component diagrams. Each component diagram has up to five small heatmaps picturing the associations discovered within or between the five data sets; see **Figure 1**. Many such associations may be found, and hence many component diagrams. In the next section, we illustrate several components and the associations within them, some of which are known while others may offer novel insights into the molecular mechanisms of obesity.

**Fig. 1.**
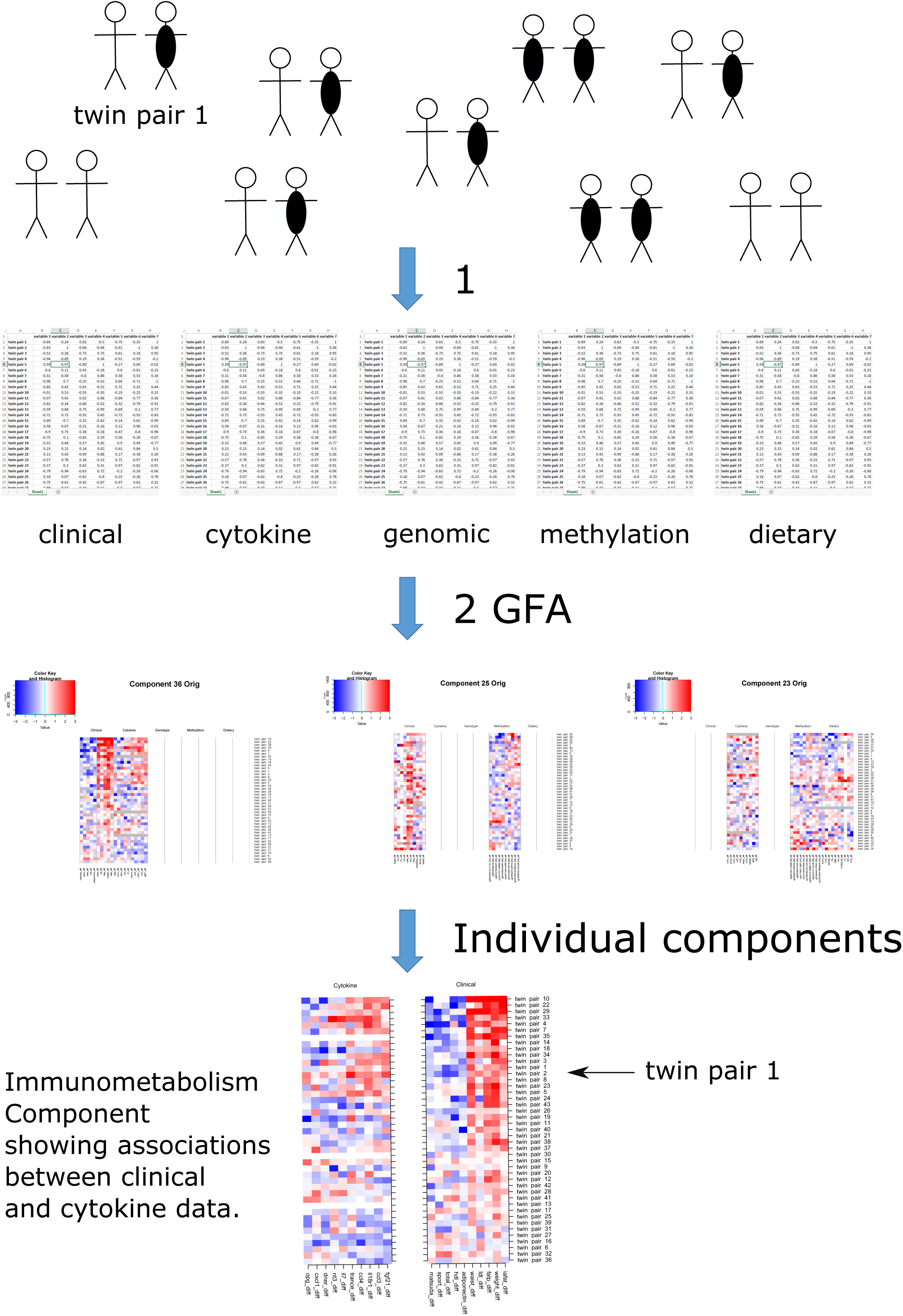
Pipeline for the GFA analysis on MZ twin pairs: (1) Clinical, cytokine, genomic, methylation and dietary data were collected from 43 young adult monozygotic twin pairs, where 25 of the twin pairs were substantially weight discordant (delta BMI > 3kg/m2). For each twin pair and each variable, the value for the leaner twin was subtracted from the value for the heavier twin. This resulted in five large data matrices comprising 42, 71, 1587, 1605 and 63 variables, respectively. (2) All five large data matrices were input into the Group Factor Analysis (GFA) computational tool, giving rise to 38 so-called component diagrams (three of which are shown in this figure). Each component diagram has up to five small heatmaps picturing the associations discovered within or between the five data sets. The magnified component is the Immunometabolism Component, also in Figure 2.

Such novel approaches that integrate multiple sources of information provide an invaluable way to discover connections at multiple molecular levels from vast amounts of data (22-24) or to predict outcomes, as in the landmark study of Zeevi and colleagues where blood parameters, dietary habits, anthropometrics, physical activity, and gut microbiota were used to predict personalized postprandial glycaemic response to real-life meals (25). So far GFA has only been taken up by the computer science community to develop the methodology further (26; 27) and, although in the literature there have been small case studies, few real applications have been previously published. These have, to our knowledge, focused on the problem of elucidating the mechanisms of action of small molecules and so the samples within the matrices have corresponded to experimental and structural information about small molecules (28).

## 2 Results

### 2.1 Machine learning meets twin studies

In our analysis, samples refer to twin pairs and the data for each twin pair corresponds to difference values for each of the variables (i.e. the value of the variable for the heavier twin minus the value for the leaner twin of each pair). The aim of the analysis is to identify drivers or consequences of change in weight, which are difficult to distinguish by this study design. The genetic data suggests (see the end of this section) that in fact most of the 43 twin pairs in our data have a genetic predisposition to obesity. Hence the analysis has the potential to elucidate why some individuals are faring better than others, despite their genetic burden. This could be invaluable knowledge for informing prevention strategies. The output of GFA applied here is 38 sets of associations between variables within and between the different data sets. As mentioned above, each such set of associations between variables is called a component and can be visualised as up to five adjacent heatmaps corresponding to the five data sets, showing how the relevant variables in each data set are associated with each other.

With the analysis producing 38 components, it is not possible here to go through all components in detail. Instead, we pick six interesting examples, choosing to include some accompanying component diagrams in the supplementary files rather than in the main body of text. We stress that many components highlight known associations, thus adding to the credibility of this approach, and we point these out for those components discussed.

In what we have called the **Immunometabolism Component (Figure 2)**, the GFA method has picked up associations between two of the five data sets, namely the clinical data and the cytokine data. These results are completely data driven and the associations of the clinical variables with obesity are all well-known (29; 30). For the twins that differ most within pair with respect to adiposity (weight, waist, fat percentage and intra-abdominal fat) as well as LDL cholesterol (seen at the top of the figure in red), the heavier twin tends to have much lower insulin sensitivity (depicted by the Matsuda index (31)), less physical activity (sport and total indices from the Baecke questionnaire (32)), HDL and adiponectin (shown by the blue colour). What may yield novel insights are the links with the cytokine data. For the twin pairs with the above profile, the heavier twin tends to have elevated values of the cytokines TRANCE, CCL4, IL18R1, CCL3 and FGF21. It has been known since the early 2000s that many immune mediators are abnormally produced or regulated in obesity, contributing to altered metabolic status (33).

**Fig. 2.**
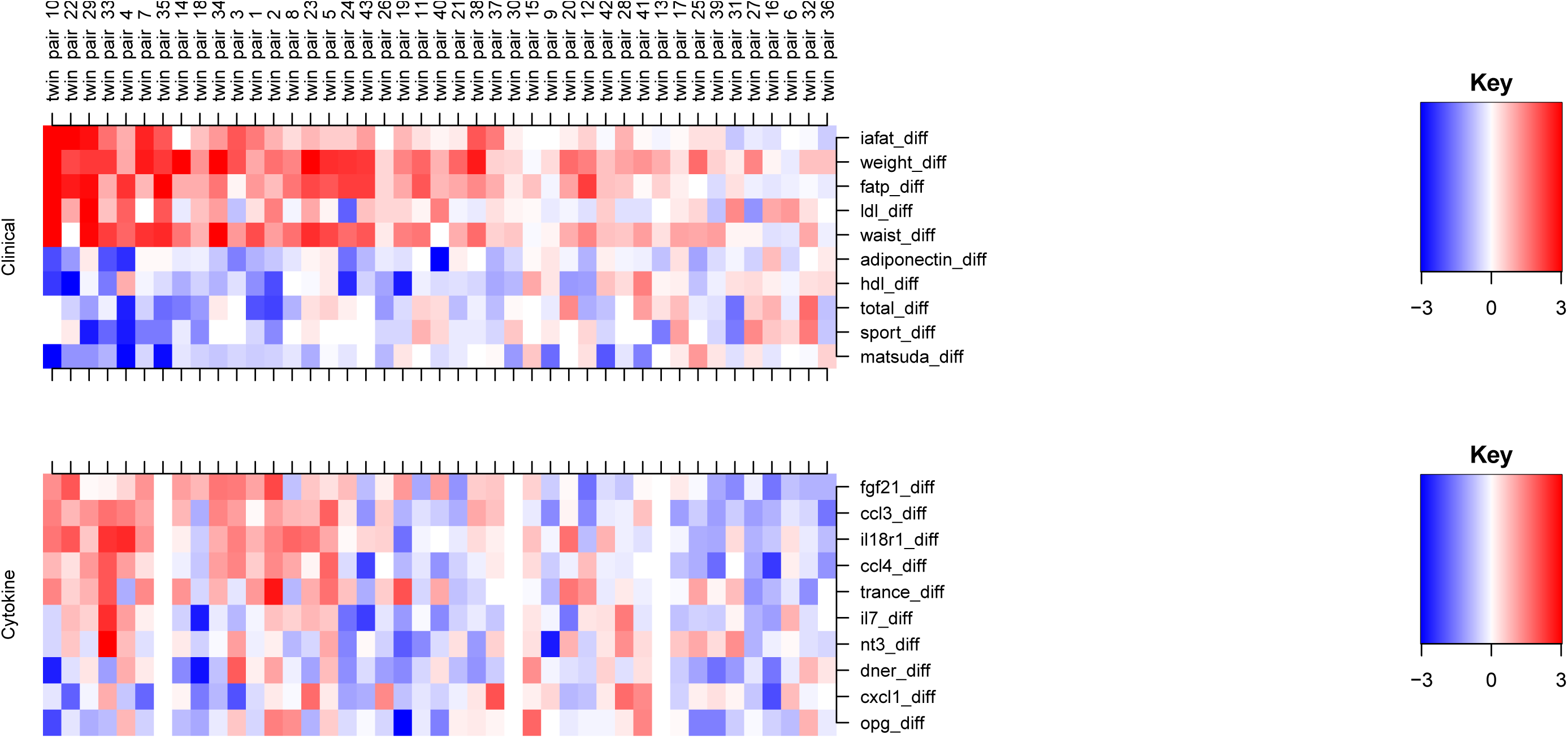
The Immunometabolism Component. The method has picked up associations between clinical data and cytokine data. The twin pairs seem to be ordered roughly by fat percentage (fatp) discordance, with the most discordant pairs at the top of the picture also having high negative HDL and adiponectin difference. Since we are mostly working with difference values, the BMI column will throughout the analyses be completely red, since the BMI for the heavier twin minus the BMI for the leaner twin will always be by design a positive value. Likewise, through all of the components certain other variables highly correlated with BMI are completely red, such as weight, subcutaneous fat, liver fat and fat percentage, and variables inversely correlated with BMI will be blue, such as adiponectin. Full description of the clinical variables: adiponectin: fasting plasma adiponectin concentration, fatp: percentage body fat, hdl: fasting plasma high density lipoprotein concentration, iafat: intra-abdominal fat volume, ldl: fasting plasma low density lipoprotein concentration, matsuda: Matsuda index, sport: sport index, total: total physical activity index, waist: waist circumference, weight: body weight.

For example, the method clearly picks up fibroblast growth factor 21 (FGF21), a hormone that is known to have elevated levels in insulin-resistant morbidities such as obesity and T2DM (34). Elevated FGF21 levels in these diseases are suspected to be signs of FGF21 resistance (35), similar to the concept of insulin resistance. FGF21 is a peptide hormone secreted by multiple tissues, most notably the liver. Indeed, we have previously shown that high values of FGF21 (measured by the enzyme-linked immunosorbent assay ELISA) are associated with high liver fat (36). Elevated levels also correlate with liver fat content in non-alcoholic fatty liver disease (37). Significantly increased FGF21 levels in circulation have been detected in patients with muscle-manifesting mitochondrial diseases, in which case most of the circulating FGF21 arises from the muscle (38). Interestingly, a single-nucleotide polymorphism (SNP) of *FGF21* – the rs838133 variant – has been identified as a genetic mechanism responsible for the sweet tooth behavioural phenotype, a trait associated with cravings for sweets and high sugar consumption (39; 40).

The other four cytokines included in this component also have some previous links to obesity, highlighting the potential of the method to offer both known and novel hypotheses on the mechanisms of immunometabolism. TRANCE (RANKL) has been proposed to link Metabolic Syndrome and osteoporosis (41). The CC chemokine family members CCL3 and CCL4 have both tens of publications where obesity is mentioned, but with debate on the mechanisms involved and function (42). Finally, although there is not much reference to IL18R1 and obesity, its ligand, IL18, has in several studies been associated with obesity, insulin resistance, hypertension and dyslipidemia (see (43) and the references therein).

A second component, the **HDL Component**, has also picked up the clinical and cytokine data sets, but this time the twin pairs seem to be ordered roughly by HDL discordance rather than weight discordance, with the twin pairs at the top of the heatmap picture (**Supplementary Figure S1**) being those for whom the heavier twin has lower levels of HDL compared to the leaner twin. This associates with the heavier twin having higher levels of cytokines CCL11, UPA, FGF19, TRANCE and SCF, again offering some potentially novel associations. Not much is known about these cytokines in relation to obesity or HDL, apart from for FGF19 which, like its related hormone FGF21 mentioned earlier, is being investigated as a pharmacological target for obesity (44). In contrast to FGF21 however, metabolic diseases exhibit reduced serum FGF19 levels (45). The simultaneous increase in serum FGF21 levels is likely a compensatory response to reduced FGF19 levels, and the two proteins concertedly maintain metabolic homeostasis (46). It is interesting then that here levels of FGF19 are increased when weight gain is accompanied by a large reduction in HDL levels. It is also interesting to note that though fat percentage difference is present in this component, its values are not exactly correlated with HDL difference.

Next we consider a component where within twin pair differences in clinical variables have been associated with methylation differences. In the **Leisure Time Physical Activity Component**, leisure time physical activity (32) seems to be associated with methylation changes independently of BMI difference (**Figure 3**). When the heavier twin partakes in less leisure activity, they also have higher levels of methylation at CpG sites on *SLC11A1, MAP7, CEBPE* and *ESR1* and lower levels of methylation at *AHRR, ZAP70, GPR15, ELMSAN1* and *GOLIM4*. We observe that for most substantially weight discordant pairs, the twin with greater leisure time activity is leaner, and we have shown elsewhere that the more active twin, even in MZ pairs, remains leaner (47-49).

**Fig. 3.**
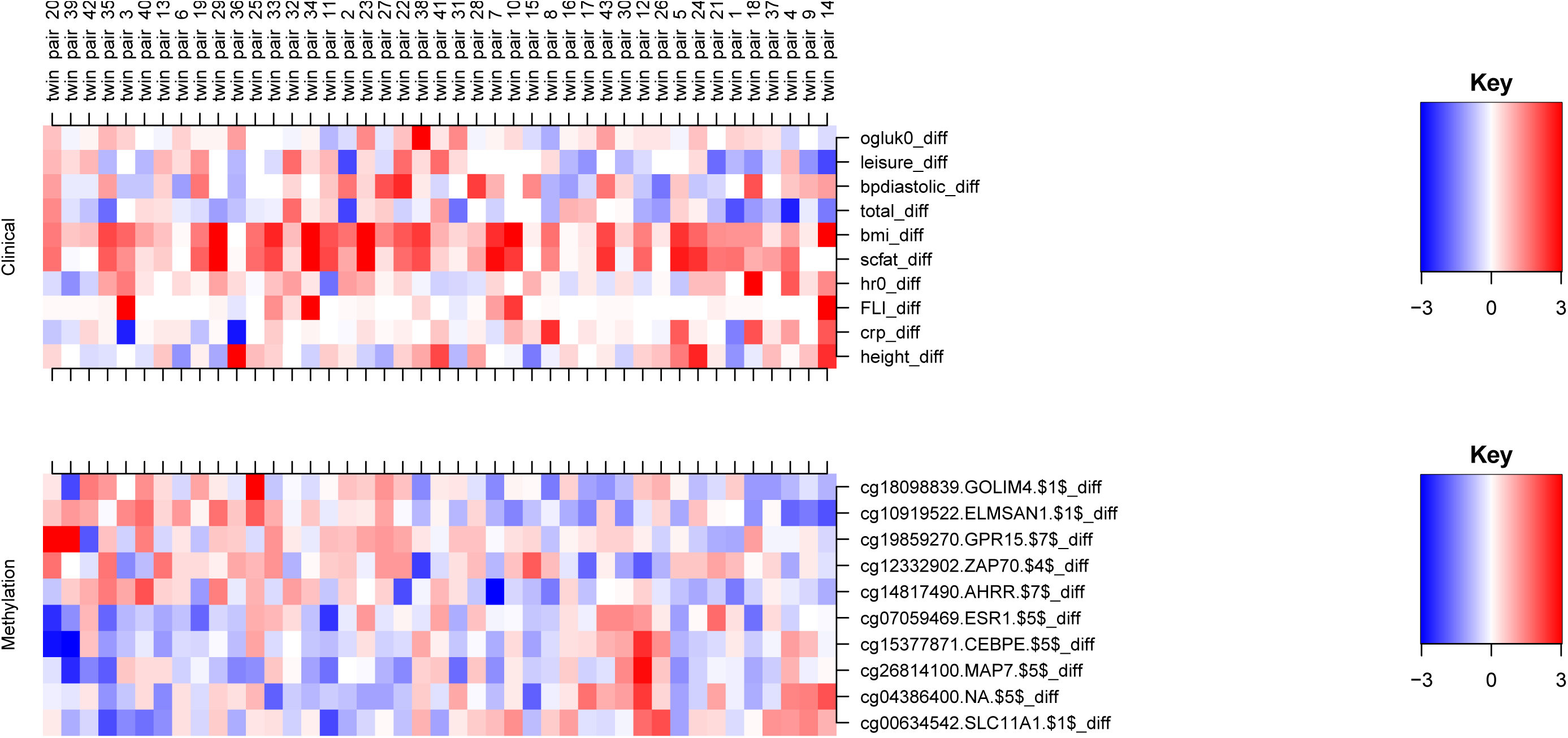
The Leisure Time Physical Activity Component. As can be seen in Table 1, the maximum within pair difference in the leisure time activity index is 1.25, which can be considered large (these index values usually have mean 3 and a range of 3.25). Full description of the clinical variables: bpdiastolic: diastolic blood pressure, bmi: body mass index, crp: C-reactive protein, FLI: Fatty Liver Index, height: height, hr0: heart rate, leisure: leisure time index, ogluk0: fasting plasma glucose concentration, scfat: subcutaneous fat volume, total: total physical activity index.

It is highly challenging to link nutrient and immune responses, let alone combining this with epigenetic alterations. Here we offer a contribution in this area. We highlight the results from three components all of which picked up associations with dietary variables.

**Table 1.**
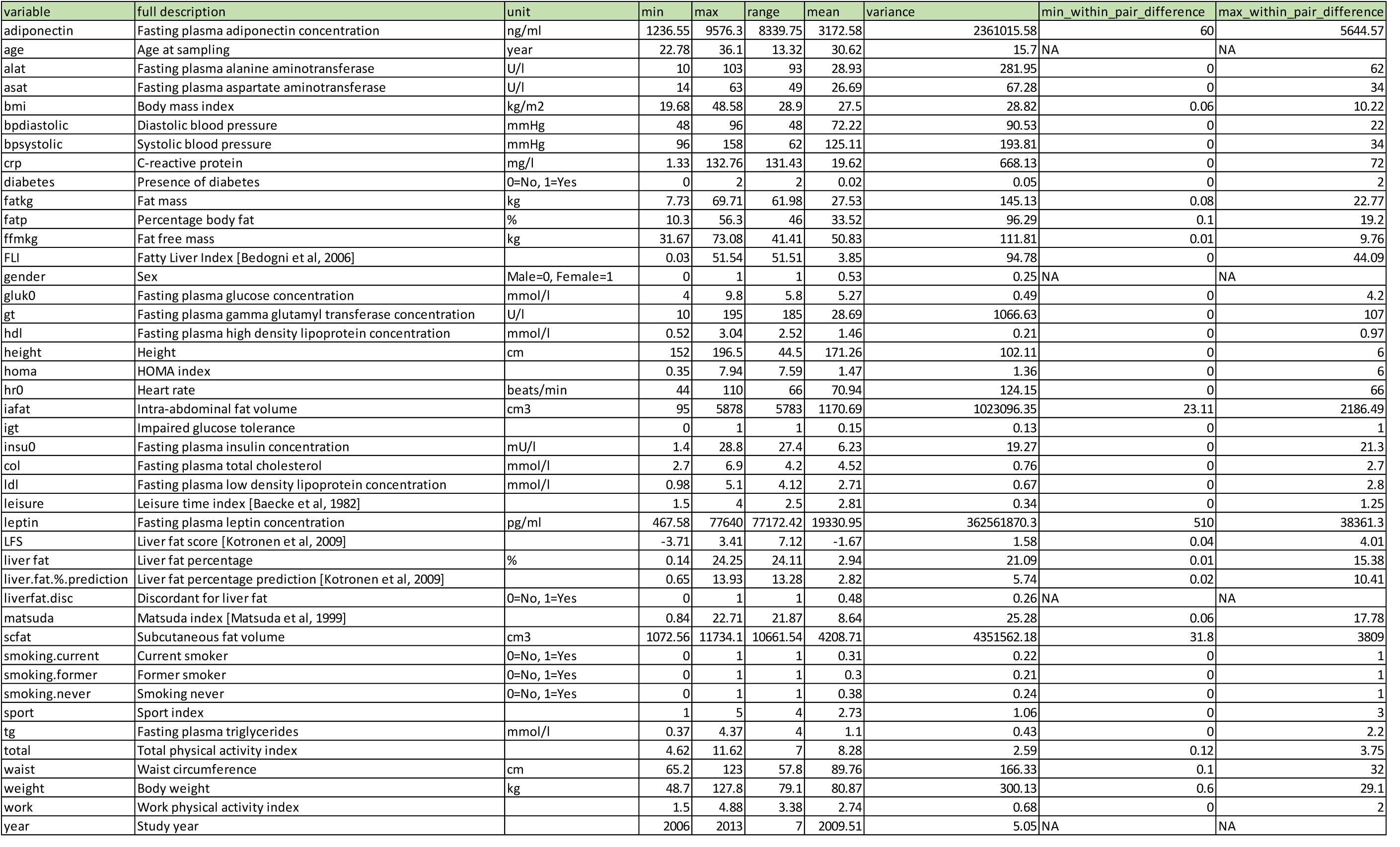
Clinical characteristics of the 43 twin pairs in our study

In the **Epigenetic Component** (**Figure 4**), we see associations between the dietary, methylation and cytokine data. When there is a clear lower consumption in the heavier twin of sucrose, vitamin D, water, fluoride and riboflavine, then there is a lower methylation of CpGs at *AHRR, INPP5D, E2F3, VMP1* and *RARA* and higher values of cytokines 4EBP1, CCL19, ENRAGE, GDNF and HGF. Although crude, this gives a glimpse into the ability of the method to highlight possible connections between these three data levels, where difference in diet is not influenced by genetics and differences in consumption are associated with differences in molecular features. All of the above genes have been linked to cytokines (mainly ILs) and inflammation in the literature and also all have associated studies on miRNAs, but it is difficult to draw any conclusions without further experiments. For the cytokines, the fit to the literature is difficult to decipher: 4EBP1 has been shown to have a protective effect in obesity in male mice (50), ENRAGE has been shown to be positively correlated with visceral fat adiposity (51), and elevated HGF levels induced by a high fat diet have been shown to have a protective role against obesity and insulin resistance (52). GDNF family members have also been implicated in obesity (53).

**Fig. 4.**
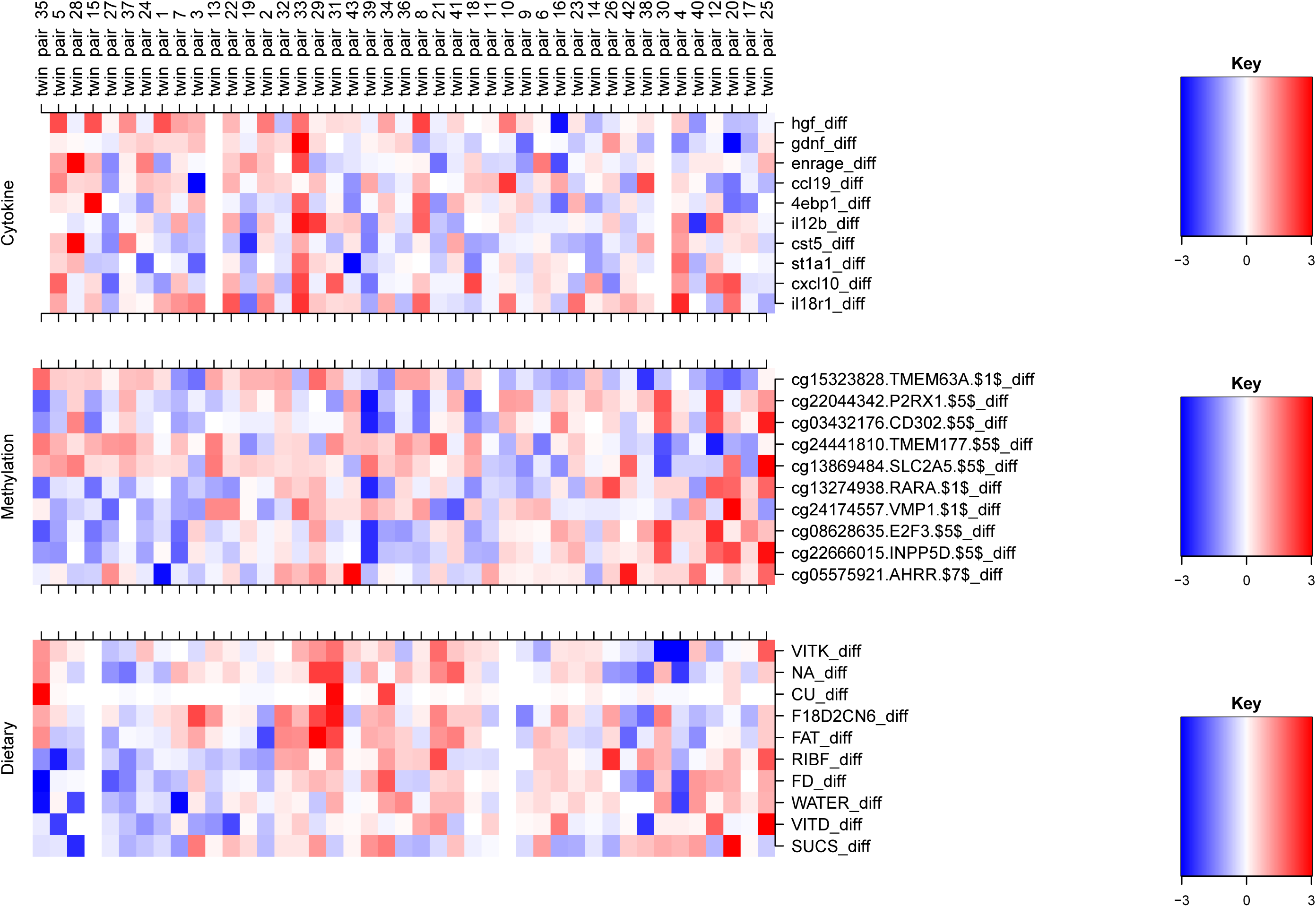
The Epigenetic Component. Full description of the dietary variables: CU: Copper, FAT: Fat, F18D2CN6: Fatty acid 18:2-n6, FD: Fluoride, NA: Sodium, RIBF: Riboflavine, SUCS: Sucrose, VITD: Vitamin D, VITK: Vitamin K, WATER: Water.

We find further links between diet and methylation in the **Sucrose Component** (**Supplementary Figure S2**). For the twin pairs at the top of the picture, the heavier twin consumes more sucrose and has increased methylation in CpGs at *CD3E, SDF4, FAM53B* and *AHRR* (a different *AHRR* CpG site to that in the epigenetics component) and decreased methylation in CpGs at *LY6G6F, C1orf127, IRX1, PPIAP3* and *MARCH11*. These individuals also consumed more carotene, vitamin C, sugar, copper and less vitamin D, molybdenum, selenium, lactose and N3 polyunsaturated fatty acids than the leaner twin in the pair. There is an apparent contradiction here between high levels of sugar and, for example, vitamin C, but it is possible that this sugar is being consumed from fruit.

It is particularly interesting that *AHRR* appears for a third time in our analysis. The methylation of this gene is commonly associated with smoking (54-56), and we do mildly see this *AHRR* - smoking association in one of our components (results not shown; the task of finding associations with smoking is hindered by the difficulty in coding into the method differences in past and current smoking status between the twins). However, there are some previous links also between *AHRR* and weight, for example in a study showing that offspring DNA methylation of *AHRR* is associated with maternal BMI and birth weight (57). Since two of our GFA components associate *AHRR* methylation with sucrose intake, we could hypothesize that sugar may also contribute to the methylation seen at *AHRR*; nutritional stimuli have been shown to contribute to *AHRR* expression (58). There is a well-established link between *AHRR/AHR* and inflammation, and dietary flavonoids and indolens, tryptophan and arachidonic acid (59-61).

In the **Starch Component** (**Supplementary Figure S3**), we have weak links between diet and clinical variables and cytokines, but do retrieve known links between lower starch consumption and lower levels of adiponectin (62). We see clearly that individuals within twin pairs who tend to eat more starch, also tend to eat more fatty acid 18:3-n3, polyunsaturated fatty acids, fluoride and carbohydrate. This may indicate use of starchy vegetables, and an overall healthier fat consumption as well. This is the only component including the gender variable, and even here the association looks quite weak. It would have been interesting to see some associations which only occurred in males or females, but we did not observe this in the current study.

Finally, the component diagrams can offer insights into the individuals in the analysis which can help inform the hypotheses drawn. With regards to the varying genetic load of the twin pairs, all appear to be susceptible to weight gain. **Supplementary Figure S4** shows five genes for which almost all of the twins have at least one risk allele, as well as interestingly another five genes associated with obesity for which hardly any of the twins have a risk allele. However, **Supplementary Figure S5** shows that the twin pairs divide into roughly two sets based on their genetic burden at five SNPs at three genes, including *FTO*. Polygenic Risk Scores (PRS) for obesity have been available for some time (63), with recent efforts producing a good prediction of those individuals at a high risk for obesity (64). In this work, we decided to retain SNP level information in order to facilitate the elucidation of molecular interactions at the genetic and other levels. However, in future work inclusion of a suitably weighted continuous PRS value could potentially distinguish differing mechanisms at high versus low PRS for obesity.

### 2.2 Results summary

The method has found many known clinical characteristics of obesity in a data-driven manner. Beyond this validation of the method, more encouraging are the suggested links between key obesity related features and mechanisms of immunometabolism. It is well known that obesity is associated with changes in the production of hormones, adipokines and cytokines (33; 65-67). A review of the growth of the field of immunometabolism and latest developments is given in two recent papers by Hotamisligil (33; 65). The work here also addresses a key question stated by Hotamisligil as to whether mechanisms can be identified that integrate nutrient and immune responses (65).

Although in the current work we are only observing associations, and so cannot claim anything regarding causality, the associations can suggest hypotheses to investigate, such as that sucrose or other dietary factors and inflammation affect methylation at *AHRR*. This could ultimately also add to efforts to develop better biomarkers of nutritional intake. We saw in the Immunometabolism Component that obesity and known related clinical variables associated with elevated levels of cytokines TRANCE, CCL4, IL18R1, CCL3 and FGF21, for which known links to obesity were discussed. In the HDL component, HDL was associated with cytokines CCL11, UPA, FGF19, TRANCE and SCF. The Leisure Time Physical Activity Component, we see that when the heavier twin partakes in less leisure time physical activity, they also have higher levels of methylation at CpG sites on *SLC11A1, MAP7, CEBPE* and *ESR1* and lower levels of methylation at *AHRR, ZAP70, GPR15, ELMSAN1* and *GOLIM4*.

Three further components focused on links between nutrient intake, immune response and methylation. We see in the Epigenetic Component that when there is a clear lower consumption in the heavier twin of sucrose, vitamin D, water, fluoride and riboflavine, then there is a lower methylation of CpGs at *AHRR, INPP5D, E2F3, VMP1* and *RARA* and higher values of cytokines 4EBP1, CCL19, ENRAGE, GDNF and HGF. In the Sucrose Component we see that when the heavier twin consumes more sucrose, carotene, vitamin C, sugar, copper and less vitamin D, molybdenum, selenium, lactose and N3 polyunsaturated fatty acids than the leaner twin in the pair, then they have increased methylation in CpGs at *CD3E, SDF4, FAM53B* and *AHRR* and decreased methylation in CpGs at *LY6G6F, C1orf127, IRX1, PPIAP3* and *MARCH11*. Finally, the Starch Component highlights the known association between lower starch consumption and lower levels of adiponectin.

Given the design of our analysis, looking at difference values within MZ twin pairs genetically predisposed to weight gain but where some individuals are faring better than others, the associations found have the potential to give hints as to why some twins of a pair differ in BMI despite their shared genetic burden. This could be invaluable knowledge for informing prevention strategies.

## 3 Discussion

What we have presented here is a relatively small scale example of the application of GFA on data from 43 MZ twin pairs. We acknowledge that this twin data is quite unique and so wish to stress that the method presented can also offer insights from non-twin data where samples are simply the measurements from individuals. In this case, the insights would be of a different form, for example “individuals of a certain genotype at given SNPs have increased consumption of x,y and z and an increase in cytokines a,b and c and decreased methylation at CpGs l, m and n”. Of course, beyond showing which variables tend to co-occur, it is difficult to know whether or how these are obesity-related without an association to the clinical variables. Hence, in this case, components including clinical variables are the most useful.

In terms of machine learning methodologies, there are two interesting future directions to explore. First, it will be interesting to explore multi-tensor factorizations such as (68) to analyse the twin-pair data sets. Tensor factorizations capture multi-way structures in the data and may reveal previously unseen patterns. In addition, it would be interesting to explore the possibility of non-linear dependencies across the data sets with kernelized matrix factorization-based approaches (69). As a further direction, machine learning innovations that allow exploring new use-cases could be explored. For example, innovations that make it easy to handle massively high-dimensional data sets such as (70) could enhance the applicability of GFA further. In addition, it could be useful to incorporate prior-biological knowledge for example in the form of pathways or functionally linked networks to supplement the model’s learning process.

There is a huge temptation to throw all possible types of data at methods such as GFA to see what novel associations can be found. We next discuss what other types of data we could include, when available.

It has previously been shown that a wide range of unfavourable alterations in the serum metabolome are associated with abdominal obesity, insulin resistance and low-grade inflammation (71) and so this would be very useful to include in future analyses. Also adding transcriptomics profiles could add clues to how genotype, DNA methylation and gene expression inter-relate (72).

There is growing evidence that the gut microbiome plays vital roles in health and disease (73) (74) and in particular that gut microbiota contribute to the regulation of adiposity and are a mediator of dietary impact on the host metabolic status, although some contradictory evidence also exists (75). Sonnenburg and Bäckhed (76) have recently reviewed evidence of how the gut microbiota can alter extraction of energy from food, generation of metabolic products, such as short-chain fatty acids, and storage of calories. Despite the likely complexity of processes involved, the broad picture seems to suggest that obesity is associated with a reduced diversity of gut microbiota (77), which would mirror the findings from macroecology suggesting that biodiversity within an ecosystem can serve as a measure of stability and robustness (78). It has also been suggested that host genetics may influence the presence of certain microbiota (15). If microbiome data were available, it would be useful to include in the GFA analysis as an additional variable indicative of the level of microbiome diversity or even a whole matrix containing microbiome or microbial taxa level data. It may also be advisable to include all putative genetic determinants of gut microbiota in the analysis. These involve genes related to diet, metabolism, olfaction and immunity (15). The number of studies into the effect of the microbiome on health is steadily increasing (79) with new methods emerging to measure its composition (80) and so we envisage many opportunities for incorporating such data into the methods in the future.

With dietary data, it is known that the unreliability of food questionnaires poses a major challenge. However, there have been recent advances in measuring dietary intake via a urine test (81) or using biomarkers in the blood (82). Such metabolic profiles could be input into the method to offer more reliable links between diet and obesity. The latest nutrition research should be considered and the breadth and resolution of dietary information adjusted accordingly in light of promising new associations. Also the role of artificial sweeteners, probiotics and emulsifiers could be evaluated if estimates of these levels were included in the data collection (73; 83-85). Finally, as some dietary compounds have been shown to influence the epigenome, inflammation and microbiota (86-89), it could be enlightening to include estimated levels of such compounds, eg resveratrol and quercetin, and some of their putative molecular targets, such as HDACs and NFkappaB, into the data (28) to begin to elucidate a more unified picture of the influence of diet.

There is evidence for an interplay between the stress system and obesity, with increased long-term cortisol levels, as measured in scalp hair, being strongly related to abdominal obesity (90). Hence adding a hair cortisol concentration measure to the variables could potentially discern stress related obesity mechanisms. Also including traditional predictors of obesity, such as parental obesity status and presence of childhood obesity, or the polygenic risk score for obesity as discussed above, might also help to distinguish different classes of obesity and their mechanisms.

It is known that genotype has a large effect on methylation and there are resources available to advise which methylation sites are influenced by the genome and which might be affected by disease relevant environmental exposures (91). In the current analysis, we decided to limit the number of SNPs and CpGs and so did not incorporate mQTL information, which would include the cis and trans effects in the analysis (72). Instead, had there been genotype-methylation associations highlighted, we planned to look up mQTL information at a later stage to help gain functional insights. In future studies, however, the mQTL genes and SNPs could also be included to see whether known associations appear and what other variables they are associated with.

## 4 Conclusions

It is now well established that machine learning coupled with good quality data is held as key to future discoveries and advances in almost every imaginable field, with investments in this area a cornerstone of the research and innovation strategies of many companies and governmental funding bodies alike. The current work contributes to efforts to bring machine learning to the field of disease prevention and in particular to obesity research.

In terms of impact of such research, discovering molecular mechanisms of disease can of course guide towards drug discovery directly. Also discovery of individual risk factors can hope to aid in disease prevention through behavioural change, although it is unclear whether informing people of genotype-based disease risk changes behaviour (92; 93), especially for those who are genetically highly susceptible to food rich environments. But at a societal level there is hope. If researchers can find incontrovertible evidence that the epidemic of obesity cannot be reversed by individual willpower alone due to the nature of the molecular mechanisms involved, then it must turn to the governments to create the environment necessary to affect the change. The interactions between the environment and the individual in the development of obesity have been acknowledged and described via a full obesity system map by the UK government in its landmark Foresight report on obesity over a decade ago (7) and there have been calls to ‘dust off’ the report and embrace even further its recommendations to adopt a ‘whole systems approach’ to tackling obesity (94). The more understanding there is of the complex molecular mechanisms involved, the greater the evidence to advocate and inform a societal effort to enable a change at an individual level.

## 5 Methods

The data from this study has been deposited in the THL Biobank and applications to access the data can be addressed to the Biobank (https://thl.fi/en/web/thl-biobank).

### 5.1 Finnish Twin Cohort

TwinFat, which is a substudy of The Finnish Twin Cohort study (FTC) (95), is designed to study obesity using an MZ co-twin control design (96; 97). The twin pairs in TwinFat were selected from two population-based twin cohorts, FinnTwin16 and FinnTwin12, comprising ten full birth cohorts of Finnish twins. Twins were included in the current study based on the availability of data for both members of a pair. All twins were free of somatic and psychiatric diseases and with a stable weight for at least three months prior to the current study. Venous blood samples were drawn in the morning after an overnight fast. Zygosity was confirmed by genotyping. All twins provided written informed consent. The protocols of the FTC TwinFat data collections were approved by the Ethics Committee of the Helsinki University Central Hospital.

### 5.2 Clinical data

Included in the analysis were 43 clinical variables. Some categorical variables (e.g. gender) are common to both twins and others (e.g. smoking status) may or may not be. For variables such as gender which are common to both twins, we include in the matrix the information from either twin (rather than difference values, which would always be zero). Gender was labelled as 0=male, 1=female. Hence in the component diagrams, red represents the female twins. Twenty twin pairs were male and 23 were female. Smoking status was recorded as never, former or current. Hence Smoking.Current is 1 if the twin currently smokes and 0 otherwise. Smoking.Current.diff is the value of Smoking.Current for the heavier twin minus that for the leaner twin (hence if the current smoking status for both twins is the same, Smoking.Current.diff will be zero). Age, Gender, Year and Liverfat Discordance were common to both twins and are input directly into the data.

We have calculated liver fat scores (LFS), and liver fat percentage prediction (98), and fatty liver index (FLI) (99), even though we actually have liver fat values measured by magnetic resonance imaging technique. The idea behind this was to see what these scores are associated with. However, we did not observe any interesting associations with liver fat or its related scores.

We also include information on physical activity (leisure time, work, sport and total physical activity indices from the Baecke questionnaire (32)), since higher physical activity level is known to be associated with lower adiposity and better metabolic health, independent of genetics (100).

Most of the twins in the study were metabolically healthy, and did not differ significantly for LDL, Homa index or insulin levels within pairs.

### 5.3 Cytokines

Included in the analysis were 71 cytokines from the Proseek Multiplex Inflammation I panel (Olink Bioscience, Uppsala, Sweden). For statistical comparison, proteins with missing data frequencies above 25% were excluded, leaving 71 proteins out of the original 92 for analysis. Protein levels are presented as normalised protein expression [NPX] values following an inter-plate control normalisation procedure.

### 5.4 Genotype information

Chip genotyping was done using Illumina Human610-Quad v1.0 B, Human670-QuadCustom v1.0 A and Illumina HumanCoreExome (12 v1.0 B, 12 v1.1 A, 24 v1.0 A, 24 v1.1 A) arrays. The algorithm for genotype calling was Illumina’s GenCall for all HumanCoreExome chip genotypes and Illuminus for 610k &670k chip genotypes. Genotype quality control was done in two batches (batch1: 610k+670k chip and batch2: HumanCoreExome chip genotypes), removing variants with call rate below 97,5% (batch1) or 95% (batch2), removing samples with call rate below 98% (batch1) or 95% (batch2), removing variants with minor allele frequency below 1% and Hardy-Weinberg Equilibrium p-value lower than 1e-06. Also samples from both batches with heterozygosity test method-of-moments F coefficient estimate value below 0.03 or higher than 0.05 were removed along with the samples which failed sex check or were among the MDS principal component analysis outliers. The total amount of genotyped autosomal variants after QC were 475637 (batch1) and 221814 (batch2). We then performed pre-phasing using Eagle v2.3 (101) and imputation with Minimac3 v2.0.1 using University of Michigan Imputation Server (102). Genotypes of both batches were imputed to 1000 Genomes Phase III reference panel (103).

Included in the analysis were 1587 SNPs and, as both twins in a pair have the same genotype, we use the information from either twin (rather than the difference values). Monozygotic twins with large BMI discordance are extremely rare. When there is discordance, it usually arises around age 16-20 but many BMI discordant pairs do not continue to be discordant when followed over time (5). This emphasizes the importance of genetic influences on weight regulation. Therefore, it is important to find the triggers to obesity and mechanisms involved for those that have a genetic predisposition.

To focus our analysis on the most important findings, we chose to include in the analyses SNPs associated with obesity and obesity related traits. An additional motivation for this choice is that, as we found in previous work (28), it is difficult to draw meaningful or actionable hypotheses from genes for which nothing is known. The main article used to choose the SNPs for this analysis was the GWAS meta-analysis of Locke et al (8). We also used SNPs retrieved from searches for BMI, liver disease, metabolic syndrome and diabetes from the NHGRI-EBI GWAS Catalog (104) as well as the SNPs from the paper of Turcot et al (105) on rare variants associated with BMI (**Supplementary Table 1**; the number in between dollar signs refers to the source from which the SNP was chosen, and is also included in the component diagrams). SNPs that are present in more than 38 pairs were removed as these are unlikely to be linked with the differences in the pairs and would only bias the model towards a different “locally” optimal solution. For each SNP, we assigned the values of no risk allele 0, one risk allele 1, and two risk alleles 2.

### 5.5 Methylation information

Methylation is one of the most stable epigenetic processes, but, on one hand, some regions are more reactive than others and, on the other hand, some marks are really stable through the lifespan, for example some of the sites associated with smoking. Twin studies have shown that DNA methylation profiles are more divergent in older twins than infant twins, and although stochasticity may have a role in this phenomenon, these findings also add support to the influence of environmental factors to the epigenome (106).

Several epigenome-wide association studies have now been conducted and have identified a panel of gene loci where methylation levels significantly differ in obese and lean individuals. We include in our analysis DNA methylation data measured on Illumina’s Infinium HumanMethylation450 BeadChip. DNA methylation measured as beta values (ranging from 0 to 1) was preprocessed using the R package methylumi (107) and normalized by beta-mixture quantile normalization (BMIQ) (108) after which the ComBat function in the R package SVA (109) was used to correct for potential batch effects. DNA methylation values of a set of pre-selected 1605 CpGs were input into the GFA method. CpG sites were selected from the recent meta-analysis of Wahl et al. (110). They show that BMI is associated with widespread changes in DNA methylation and genetic association analyses demonstrate that the alterations in DNA methylation are predominantly the consequence of adiposity, rather than the cause. We also include CpGs associated with elevated liver fat (19), CpGs whose methylation has been previously shown to differ in the adipose tissue of BMI-discordant MZ twin pairs (111), smoking-associated CpGs (112), and CpG sites that have been associated with weight loss (113) (**Supplementary Table 2**).

### 5.6 Dietary data

Included in the analysis were 63 dietary variables (**Supplementary Table 3**). Total energy and macronutrient intake were assessed with 3-day food records. Subjects were given clear oral and written instructions by a registered dietician on how to keep the food record (two working days and one non-working day) and they were encouraged to keep their usual eating patterns and to estimate the amounts of all foods and drinks using household measures. The conversion of data from the records into nutrient values was performed by a dietician using the program DIET32, which incorporates the national Finnish database for food composition (Fineli ^R^). The nutritional composition of new ready-prepared meals that were not included in the DIET32 program was obtained from the manufacturers. Daily energy intake is expressed in kilocalories (kcal) and macronutrient intakes are expressed as percentages of total energy intake. Results are presented as the mean ± SD of the 3 days (114). Information on habitual diet was estimated using a qualitative food-frequency questionnaire (FFQ) incorporating 52 food and non-alcoholic beverage items that are common in the Finnish diet (115).

### 5.7 Group Factor Analysis

Group Factor Analysis (GFA) is formulated as a method to identify statistical dependencies between multiple data sets. The method learns a joint integrated model of the datasets with matched samples (i.e. having a common set of samples), to extract meaningful and interpretable information (26; 116; 117). GFA has been successfully used for identifying structural properties predictive of drug responses (23), cross-organism toxicogenomics (118) as well as highly accurate drug response predictions (27).

Specifically, GFA takes a set of matrices **X**^(m)^, where m = 1…M, and identifies patterns of statistical dependencies across all of them. The model learns these statistical dependencies in a data-driven fashion automatically identifying the type and amount of dependencies. Therefore, GFA learns a low-dimensional space **Z** (of K components) that represent the variation patterns across all data sets. A component can be active in one or more data sets, meaning that it captures the relationships between those particular data sets. This is achieved by modelling the total variation of all of the datasets while inducing structured sparsity. This characteristic allows GFA to automatically identify dependency patterns that are shared across any subset of the data sets, in a truly data-driven fashion.

Formally, GFA is defined in a Bayesian formulation for data sets 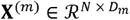, where m = 1… M, having N matched samples and D_m_ dimensions, as a product of the Gaussian latent variables Z ∈ ℛ ^*N* × *K*^ and projection weights 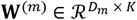:

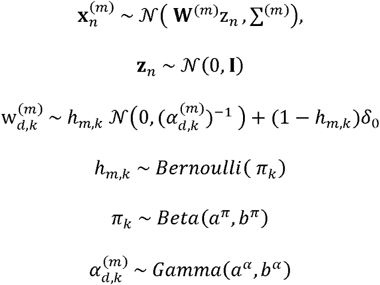

Here, ∑^(m)^ is a diagonal noise covariance matrix. GFA induces component-wise sparsity on the projections 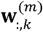 using a Beta-Bernoulli distribution. As a result, the projection weights 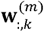 for a component k are active for the one or more data sets (m), capturing dependencies specific to one data set or shared between data sets.

The R package GFA version 1.0.3 (119), is used to model the five data sets, dietary, clinical, genomic, methylation and cytokine. In the genomic data, SNPs with values in three or less samples and more than 38 (43-5) samples were removed as outliers. In the clinical data set, the categorical variable smoking [never, former, current] was one-hot encoded to form binary representation. Additionally, all variables with more than 50% of values missing were left out of the matrices. As a result, for the N=43 twin pair samples, the clinical data contained D_1_ = 42 variables, cytokine D_2_ = 71, genotype D_3_ = 1587, methylation D_4_ = 1605 and dietary D_5_= 63. All data sets except genotype were scaled to unit variance.

The model, implemented using Gibbs sampling, was run with K = 40 components which was deemed large enough due to the presence of empty of components, as recommended by Virtanen et. al. (116). In order to model the large amount of noise in the data, we used an informative noise prior with a priori noise variation set to 1/3 (26). Finally, a total of 2000 sampling iterations were run, with the first 90% corresponding to burnin while the last 10% representing the posterior.

GFA found 38 components to model the five data sets. Components active in two or more data sets, such as clinical and cytokine, represent variation common to the two data sets. Components specific to a data set, for example genotype, represent variation patterns that are consistent within the genotype but not correlated with other data sets. Both types of components represent interesting relationships in our case and are examined in this study.

## Supplementary Files

**Supplementary Figure S1: The HDL Component** As can be seen in Table 1, the largest within pair difference in HDL is 0.97, where a difference of more than 0.5 is usually considered to be large. Full description of the clinical variables: bpsystolic: systolic blood pressure, col: fasting plasma total cholesterol, fatp: percentage body fat, hdl: fasting plasma high density lipoprotein concentration, homa: HOMA index, iafat: intra-abdominal fat volume, liver.fat: liver fat percentage, liver.fat.%.prediction: liver fat percentage prediction, matsuda: Matsuda index, smoking.current: current smoker.

**Supplementary Figure S2: The Sucrose Component** Full description of the dietary variables: CU: Copper, CAROT: Carotene, FAPUN3: N3 polyunsaturated fatty acids, LACS: Lactose, MO: Molybdenum, SE: Selenium, SUGAR: Sugar, SUCS: Sucrose, VITC: Vitamin C, VITD: Vitamin D.

**Supplementary Figure S3: The Starch Component** Full description of the dietary variables: CHO: Carbohydrate, CU: Copper, FAPU: Polyunsaturated fatty acids, FD: Fluoride, F18D3N3: Fatty acid 18:3-n3, RETOL: Retinol, r44m: Fresh vegetables, STARCH: Starch, THIA: Thiamine, VITA: Vitamin A. Full description of the clinical variables: adiponectin: fasting plasma adiponectin concentration, diabetes: presence of diabetes, ffmkg: fat free mass, gender: sex, gt: fasting plasma gamma glutamyl transferase concentration, liver.fat: liver fat percentage, matsuda: Matsuda index, ogluk0: fasting plasma glucose concentration, smoking.never: smoking never, tg: fasting plasma triglycerides.

**Supplementary Figure S4: Genotype Component 1** Blue means that the twins in the pair (who share the same genotype) carry no disease allele at the particular SNP, white means that they carry one disease allele and red that they carry two copies of the disease allele.

**Supplementary Figure S5: Genotype Component 2** The twin pairs seem to divide roughly into two groups with the twin pairs at the bottom of the figure almost all having either one or two copies of the disease allele (white or red) in the SNPs in the left hand column and the twin pairs at the top of the figure mostly not carrying a disease allele in these SNPs (blue). There was no clear way to distinguish these two groups for example in terms of the level of BMI discordance.

**Supplementary Table 1: Table of SNPs used in the analysis and the sources from which they were chosen** To focus our analysis on the most important findings, we chose to include in the analyses SNPs associated with obesity and obesity related traits. An additional motivation for this choice is that, as we found in previous work (28), it is difficult to draw meaningful or actionable hypotheses from genes for which nothing is known. The main article used to choose the SNPs for this analysis was the GWAS meta-analysis of Locke et al (8). We also used SNPs retrieved from searches for BMI, liver disease, metabolic syndrome and diabetes from the NHGRI-EBI GWAS Catalog (104) as well as the SNPs from the paper of Turcot et al (105) on rare variants associated with BMI. The number in between dollar signs links to the source from which each SNP was chosen, and is also included in the component diagrams.

**Supplementary Table 2: Table of CpGs used in the analysis and the sources from which they were chosen** CpG sites were selected from the recent meta-analysis of Wahl et al. (110). They show that BMI is associated with widespread changes in DNA methylation and genetic association analyses demonstrate that the alterations in DNA methylation are predominantly the consequence of adiposity, rather than the cause. We also include CpGs associated with elevated liver fat (19), CpGs whose methylation has been previously shown to differ in the adipose tissue of BMI-discordant MZ twin pairs (111), smoking-associated CpGs (112), and CpG sites that have been associated with weight loss (113). The number in between dollar signs links to the source from which each CpG was chosen, and is also included in the component diagrams.

**Supplementary Table 3: Table of the 63 dietary variables used in the analysis**

## Acknowledgements

MK would like to thank Leonie Bogl for very interesting and useful discussions on the dietary data in the TwinFat study; JK has been supported by the Academy of Finland (grants 308248 &312073). Funding sources for KHP are the Academy of Finland (grants 272376, 266286, 314383, 315035), Finnish Medical Foundation, Finnish Diabetes Research Foundation, Novo Nordisk Foundation, Gyllenberg Foundation, Sigrid Juselius Foundation, Helsinki University Hospital Research Funds, Government Research Funds and University of Helsinki. MO has been supported by the Academy of Finland (grant 297908), Sigrid Juselius Foundation, and University of Helsinki. The funders had no role in planning study design, data collection, and analysis, decision to publish, or preparation of the article.

